# Disentangling the association of hydroxychloroquine treatment with mortality in Covid-19 hospitalized patients through Hierarchical Clustering

**DOI:** 10.1101/2021.01.27.21250238

**Authors:** Licia Iacoviello, The COVID-19 RISK and Treatments (CORIST) Collaboration

## Abstract

The efficacy of hydroxychloroquine (HCQ) in treating SARS-CoV-2 infection is harshly debated, with observational and intervention studies reporting contrasting results.

To clarify the role of HCQ in Covid-19 patients, we carried out a retrospective observational study of 4,396 unselected patients hospitalized for Covid-19 in Italy (February-May 2020). Patients’ characteristics were collected at entry, including age, sex, obesity, smoking status, blood parameters, history of diabetes, cancer, cardiovascular and chronic pulmonary diseases and medications in use. These were used to identify subtypes of patients with similar characteristics through hierarchical clustering based on Gower distance. Using multivariable Cox regressions, these clusters were then tested for association with mortality and modification of effect by treatment with HCQ.

We identified two clusters, one of 3,913 younger patients with lower circulating inflammation levels and better renal function, and one of 483 generally older and more comorbid subjects, more prevalently men and smokers. The latter group was at increased death risk adjusted by HCQ (HR[CI95%] = 3.80[3.08-4.67]), while HCQ showed an independent inverse association (0.51[0.43-0.61]), as well as a significant influence of cluster*HCQ interaction (p<0.001). This was driven by a differential association of HCQ with mortality between the high (0.89[0.65-1.22]) and the low risk cluster (0.46[0.39-0.54]). These effects survived adjustments for additional medications in use and were concordant with associations with disease severity and outcome.

These findings suggest a particularly beneficial effect of HCQ within low risk Covid-19 patients and may contribute clarifying the current controversy on HCQ efficacy in Covid-19 treatment.

## Introduction

Hydroxychloroquine (HCQ) is an antimalarial drug suggested to be effective in inhibiting Severe Acute Respiratory Syndrome Coronavirus 2 (SARS-Cov-2) replication in vitro [1,2]. Indeed, HCQ is characterized by anti-viral, anti-inflammatory, and anti-thrombotic actions, contrasting the main disruptive effects of SARS-CoV-2 infection on the organism [3]. For this reason, it has been heavily used in treating patients affected by SARS-CoV-2 infection related disease (commonly known as Covid-19), especially in the first phases of the current pandemics, when Covid-19 was quite unknown [4].

Despite these elements and initial suggestive evidence of efficacy based on daily clinical practice, in the last months the potential benefit of HCQ for Covid-19 patients has been harshly debated [3,5]. In particular, evidence supporting protective effects from observational studies [6–13] was in contrast with that suggesting no effect at all by recent randomized clinical trials (RCTs) [14–18]. More recently, a meta-analysis combining both RCTs and observational studies over more than 44,000 patients supported a protective effect of HCQ, driven by the findings of observational studies [5]. A potential explanation for this discrepancy may be due to the usually high dosage administered in RCTs (800 mg/day), compared to lower dosages reported in observational studies supporting HCQ efficacy (≤400 mg/day), as hypothesized elsewhere [5]. As an alternative explanation, it is likely that the efficacy of HCQ treatment for Covid-19 may vary across patients and is influenced by subtypes of the disease, which in turn is largely dependent on patients’ characteristics and their non-linear combinations [19]. In this “personalized medicine” view, response to HCQ may not be the same across all patients of the same age, or with similar circulating inflammation levels. In order to identify these combinations, the use of big data like Electronic Health Records (EHRs) and of machine learning (ML) algorithms to interpret hidden HCQ response patterns is of fundamental importance. Notwithstanding it, to our knowledge only one study attempted so far a similar approach through the application of a supervised ML technique (gradient boosting), to identify those patients with likely beneficial effects of HCQ treatment. Interestingly, authors reported a reduction of in-hospital mortality within patients treated with HCQ, which was even more pronounced within those patients predicted to benefit most from the drug, in line with expectations [19].

Here, we attempted a personalized Covid-19 patient characterization to better disentangle the beneficial effects of HCQ previously reported within the COVID-19 RISK and Treatments (CORIST) study, a large retrospective cohort of patients hospitalized for SARS-CoV-2 infection in Italy [13]. This approach consisted of i) identifying the existence of subtypes of Covid-19 patients through an unsupervised ML algorithm - hierarchical clustering - comparing their characteristics and their clinical risks, ii) testing the resulting patients’ clusters for association with mortality and modification of effect by treatment with HCQ and iii) analyzing potential interactions between clusters and HCQ use. This approach represents a prominent example of how personalized medicine may support clinicians in Covid-19 treatment.

## Methods

### Analyzed cohort

The COVID-19 RISK and Treatments (CORIST) study includes 4,396 patients hospitalized for SARS-Cov-2 infection in 35 hospitals across Italy, between February 2020 and May 2020. Molecular diagnosis of SARS-CoV-2 infection was based on polymerase chain reaction (PCR) of viral DNA extracted and amplified from nasopharyngeal swabs. Within each participating hospital, clinical data were abstracted at one-time point from electronic medical records or charts, and collected using either a centrally-designed electronic worksheet or a centralized web-based database. Collected data included patients’ demographics, laboratory tests, medications in use, history of disease and prescribed pharmacological therapy for Covid-19 treatment. For each participant, the study index date was defined as the date of hospital admission, while the study end point was death. Follow-up time was computed as the time between the index date and death, or alternatively between the index date and the date of discharge, applying right-censoring. Further details on the study are reported elsewhere [13,20,21].

### Statistical Analyses

#### Cluster analysis

All analyses were carried out in R 4.0.2 [22]. We applied a hierarchical clustering analysis on Covid-19 patients using their main clinical, lifestyles and socio-demographic characteristics, which were suggested as the most influential on mortality risk by previous studies in the field [21,23,24]. These included age (years), sex, glomerular filtration rate (eGFR, mL/min/1.73 m^2^) and high-sensitivity plasma C-reactive protein levels (mg/L) at in-hospital admission, obesity (body mass index (BMI) ≥ 30 kg/m_^2^_), hypertension (Yes/No), smoking status (never/previous/current smoker), as well as history of myocardial infarction, heart failure, chronic pulmonary disease, cancer and diabetes (Yes/No). Missing data were imputed through a k-Nearest Neighbor approach, implemented in the *knn()* function of the VIM package (with k=10) [25]. CRP was transformed on the natural logarithm scale to reduce skewness and all the continuous variables were normalized through the *normalize()* function in keras (see URLs).

Cluster analyses were then performed on the 4,396 patients, using the variables specified above, through the cluster package (see URLs). First, we computed a dissimilarity matrix based on Gower pairwise distance (Figure S1), through the *daisy()* function. Gower distance is a parameter in the [0;1] range representing the average of partial dissimilarities across individuals (the higher the distance, the more dissimilarities for a given pair of subjects) [26]. Second, we performed hierarchical clustering through the *hclust()* function applied to the Gower distance matrix computed above, which separated subjects based on their degree of pairwise dissimilarity, both from lowest to highest (agglomerative clustering) and from highest to lowest (divisive clustering). Third, we determined the appropriate number of clusters for patient classification, based on the Average Silhouette method (Figure S2). This computes the number of clusters which maximizes the average silhouette width, a measure of the quality of a clustering indicating how well each object lies within its cluster [27]. This method, applied through the *fviz_nbclust()* function of the factoextra package (See URLs), computed k=2 as the optimal number of clusters. Finally, each patient was assigned to one of the two clusters determined above, through application of the *cutree()* function (cluster package) to the results of the cluster analysis previously carried out.

#### Comparison of clusters

First, we compared the classifications made through agglomerative and hierarchical clustering, which revealed high consistency (Odds Ratio = 34.0 [26.7-43.7], Fisher Exact Test p-value < 10^−15^). In light of this homogeneity of classification and since divisive clustering has been reported to be more accurate and robust [28], all the subsequent analyses were performed on cluster classification identified through the latter approach.

The two clusters of patients identified were then compared for all anamnestic variables mentioned above, through Fisher’s Exact Test (for binary variables), Chi-squared test (for non-binary categorical variables), and through Student’s t test or Wilcoxon Rank Sum tests (for continuous variables meeting and not meeting parametric assumptions, respectively). Similarly, we compared Covid-19 disease severity, classified by recruiting centers in asymptomatic/mild, non-severe pneumonia, severe pneumonia and acute respiratory distress syndrome (ARDS). Moreover, we compared the use of six common drugs for Covid-19 treatment between the two clusters, including hydroxychloroquine, antihypertensive drugs, anti-interleukin-6 antibody, antivirals (Remdesivir, Lopinavir, Darunavir) and corticosteroids. These were reported as binary variables (Yes/No) and were therefore compared with clusters through Fisher Exact Tests.

#### Survival analyses

Once the clusters were characterized, we modelled incident mortality risk as a function of patients clusters and use of HCQ through Cox Proportional Hazards (PH) models, using the *cox*.*zph()* function of the survival package (see URLs) [29]. Only patients with complete information needed in each model were included in the analysis (case-complete approach, see below). A preliminary check of the basic Cox PH assumptions revealed no influential observations based on dfbeta residuals (Figure S3a), while Schoenfeld residuals tests revealed a statistical violation of the proportionality of hazards assumption, although these did not show any evident trend at a visual inspection (Figure S3b). For this reason, we carried out Cox PH models both with and without including an interaction term with time-to-event, a strategy commonly used to overcome this violation [30]. Incrementally adjusted models were analyzed: i) a crude model including only patients’ clusters (Model 1; N = 4,319); ii) a model testing additive influence of clusters and HCQ use (Model 2: Model 1 + HCQ; N = 4,212) and iii) a model testing both additive and synergistic influence of clusters and HCQ use (Model 3: Model 2 + clusters*HCQ; N = 4,212). Additional sensitivity analyses were carried out to rule out potential confounding effects of additional drugs in use for Covid-19 treatment (Model 4: Model 3 + other drugs). Risk estimates were computed as hazard ratios (HR) with 95% confidence intervals (95% CI) of dying, and HR with p-values below α = 0.05 were considered significant. To quantify the potential for unmeasured confounding effects, the E-value was calculated for all the HRs observed in Model 3, as described in [31](see URLs). This represents the minimum association required for a potential unmeasured confounder with both the exposure and the outcome to explain away the observed association between the latter. In other words, the higher the E-value, the harder it is to attribute an association to an unmeasured covariate [32].

#### Associations with additional endpoints

To better evaluate the associations of Covid-19 patients clusters and HCQ use with negative outcomes other than death, we built a composite endpoint based on the occurrence of at least one of the following outcomes: in-hospital death, access to intensive care unit during hospitalization or severe disease manifestation (either severe pneumonia or ARDS). In this case, the resulting binary variable was assigned a value of 1. Conversely, the variable got “0” value if one of the following alternative conditions applied: i) none of the above mentioned outcomes was verified; ii) a patient survived without recurring to intensive cares or iii) without showing severe manifestations of the disease. Six patients with missing values on survival were removed. Then, we modelled the risk of manifesting a bad outcome through a logistic regression (*glm()* function in R), modelling both additive and interactive models of Covid-19 patients cluster and HCQ use, as above. This analysis was motivated by the fact that the curse of disease often differs across patients, e.g. with some subjects with less severe forms suddenly worsening their conditions until death and others having severe manifestations but still surviving, possibly thanks to intensive cares. Therefore, a composite outcome variable represented a robust way to measure potential risk/protective effects of patients’ clusters and HCQ use.

## Results

While both agglomerative and divisive clustering approaches were developed, all statistical analyses presented below are based on clusters identified through the latter approach, since this showed a high homogeneity with the results of agglomerative clustering (see Methods section) and divisive clustering has been reported to be more accurate and robust [28,30].

### Characteristics of the clusters

We identified two clusters of Covid-19 patients (with N = 3,913 and 483, Figure 1). The larger cluster was younger (mean (SD) age: 65.2 (15.6) vs 77.9 (9.2) years; t-test = -25.9, p < 10^−15^), with better kidney function (eGFR: 77.9 (26.9) vs 52.6 (25.6) mL/min/1.73 m^2^; t-test = 20.3, p < 10^−15^) and lower circulating inflammation levels (CRP: 34.6 (62.4) vs 36.5 (58.6) mg/L; Wilcoxon-test = 847,660, p = 2.5×10^−14^) (Figure 2a-c). Conversely, BMI did not show strong differences between the two clusters (28.0 (4.2) vs 27.5 (4.2) Kg/m^2^; t-test = 2.3, p = 0.02; Figure 2d). Moreover, patients belonging to the larger cluster were less frequently men (60% vs 75%) and smokers (11.5% vs 19.5%) and showed a lower prevalence of chronic health conditions like myocardial infarction, heart failure, diabetes, hypertension, cancer and lung disease (all p<0.0001), while no significant difference was observed in the prevalence of obesity (Table 1).

**Table 1.** Comparison of main categorical variables between the two clusters identified.

| **Category (%)** | **Cluster 1 – low risk**<br>N = 3,913 | **Cluster 2 – high risk**<br>N = 483 | **p for difference** |
| --- | --- | --- | --- |
| Men | 2,346 (60.0%) | 362 (74.9%) | $6 \times 10^{-11}$ |
| Smoke |  |  |  |
| Current smokers | 450 (11.5%) | 94 (19.5%) | $< 10^{-15}$ |
| Previous smokers | 268 (6.8%) | 94 (25.3%) |  |
| Obesity<br>(BMI $\geq 30$ Kg/m<sup>2</sup>) | 546 (13.9%) | 64 (13.3%) | 0.73 |
| Myocardial Infarction | 127 (3.2%) | 335 (69.4%) | $< 10^{-15}$ |
| Heart Failure | 171 (4.4%) | 315 (65.2%) | $< 10^{-15}$ |
| Diabetes | 621 (15.9%) | 276 (57.1%) | $< 10^{-15}$ |
| Hypertension | 1,828 (46.7%) | 453 (93.8%) | $< 10^{-15}$ |
| Cancer | 392 (10.0%) | 89 (18.4%) | $2 \times 10^{-07}$ |
| Lung Disease | 415 (10.6%) | 207 (42.8%) | $< 10^{-15}$ |

P for difference resulting from comparison of the clusters – through Fisher’s Exact Test (for binary variables) or Chi-squared test (for non-binary categorical variables, i.e. smoke) – are reported, along with absolute and % frequency of each condition within each cluster.

**Figure 1.**
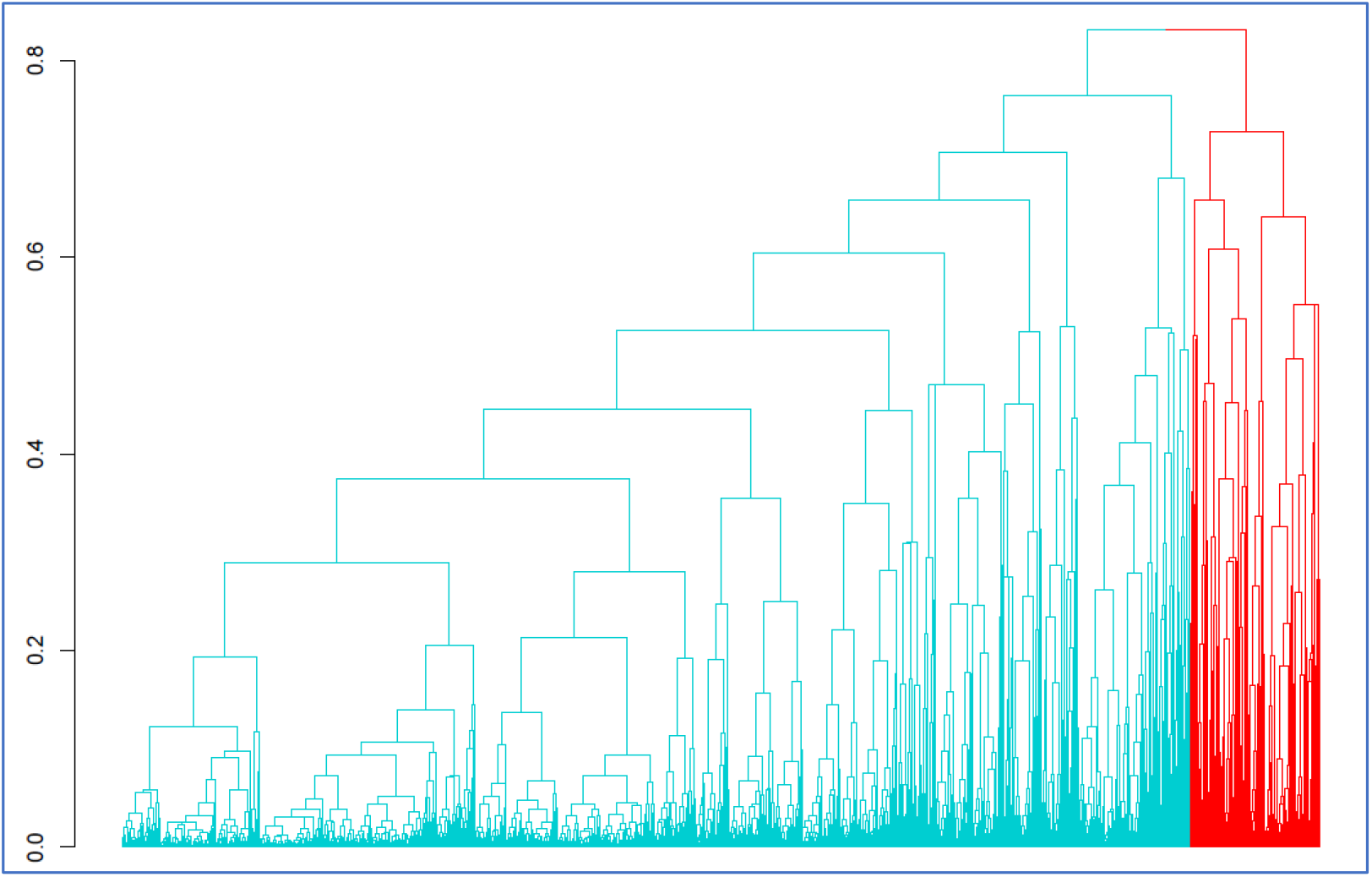
Hierarchical divisive clustering of Covid-19 hospitalized patients. Two main clusters of patients were identified, with N= 3,913 (green) and 483 (red) respectively.

**Figure 2.**
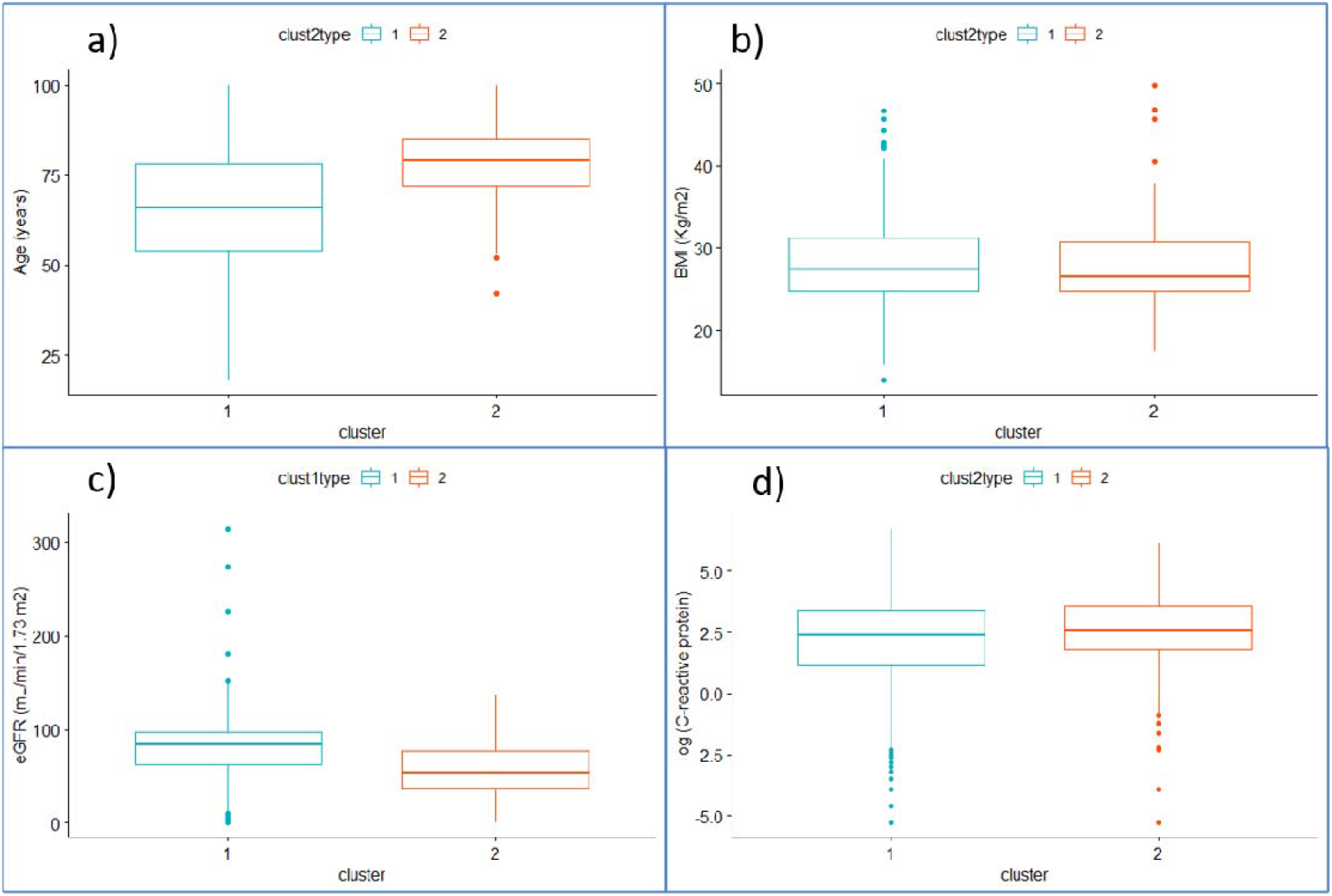
Characteristics of sample according to the two clusters identified. Comparison of the continuous variables used for hierarchical clustering - including **a)** age (years), **b)** BMI (Kg/m^2^), **c)** eGFR (mL/min/1.73 m^2^) and **d)** C-reactive protein plasma levels (mg/L, logscale)between the two clusters of Covid-19 patients identified, namely the low (green) and the high risk (red) cluster.

Clusters were also associated with severe Covid-19 disease manifestations, with 65.8% of patients in the smaller cluster presenting with either severe pneumonia or ARDS, compared to 45.9% in the larger cluster (Chi-squared = 76.4, p < 10^−15^; Table S1). For the characteristics mentioned above, the large and small clusters will be hereafter named as “low risk” and “high risk” cluster. When we compared the use of specific drugs, in the high risk cluster we observed a less frequent use of HCQ (p < 0.001) and of Lopinavir/Darunavir (p < 0.05) and a more frequent use of corticosteroids and antihypertensive medications (p < 0.001), compared to the low risk cluster (Table S2).

### Combined influence of clusters and HCQ use on mortality risk

In Cox PH regressions modelling mortality risk as a function of clusters (Model 1), we analyzed 4,319 patients with a case-complete approach, with a total of 799 deaths and a total follow-up 73,924 person-days (median 13 days). In this model, patients belonging to the high risk cluster showed a significant increase of incident mortality risk, compared to those of the low risk cluster (HR [CI] = 3.81 [3.12-4.65]; Table 2). This association remained stable in a Cox regression modelling additive effects of clusters and HCQ use (Model 2: N = 4,212, 743 events, total follow-up 72,239 person-days, median 14 days). Indeed, the high risk cluster was associated with a significant increase of mortality (3.80 [3.08-4.67]), while HCQ use was associated with a significant independent reduction (0.51 [0.43-0.61]). When we modelled additive and interactive associations of clusters and HCQ in a single model (Model 3), we observed a substantially stable protective association of HCQ (0.46 [0.38-0.55]), a reduced but still significant direct association of the “high risk” cluster (2.45 [1.69-3.54]), and a significant association of the cluster*HCQ interaction term with incident mortality (p = 2.1×10^−4^). This was driven by a differential association of HCQ use within the different clusters, since this was associated with a notable reduction of mortality risk in the low risk cluster (HR [CI] = 0.46 [0.39-0.54], p < 10^−15^) and with a milder non-significant reduction in the high risk cluster (0.89 [0.65-1.22], p = 0.47). The above mentioned associations were quite robust against potential unmeasured cofounding effects, with E-values of 3.1, 2.8 and 2.5 for the associations of mortality risk with cluster 2, HCQ use and cluster*HCQ interaction in Model 3. Moreover, these associations remained substantially stable in Cox PH models including additional drugs in use (Table 2, Model 4), as well as in those including an interaction term with time (Table S3).

**Table 2.** Results of Cox PH regressions modelling incident mortality risk.

| **Model** | **N**<br>**(deaths)** | **Cluster 2 vs 1** | **HCQ**<br>**Yes vs no** | **Cluster\*HCQ** |
| --- | --- | --- | --- | --- |
| Model 1: Death □ Cluster | 4,319<br>(799) | **3.81**<br>**[3.12-4.65]**<br>**(< 10<sup>-15</sup>)** | - | - |
| Model 2: Death □ Cluster + HCQ | 4,212<br>(743) | **3.80**<br>**[3.08-4.67]**<br>**(< 10<sup>-15</sup>)** | **0.51**<br>**[0.43-0.61]**<br>**(8.8×10<sup>-15</sup>)** | - |
| Model 3: Death □ Cluster + HCQ<br>+ Cluster*HCQ | 4,212<br>(743) | **2.45**<br>**[1.69-3.54]**<br>**(4.9×10<sup>-4</sup>)** | **0.46**<br>**[0.38-0.55]**<br>**(< 10<sup>-15</sup>)** | **1.90**<br>**[1.21-2.96]**<br>**(2.1×10<sup>-4</sup>)** |
| Model 4: Death □ Cluster + HCQ<br>+ Cluster*HCQ + Other Drugs | 3,736<br>(664) | **1.65**<br>**[1.20-2.26]**<br>**(2.2×10<sup>-3</sup>)** | **0.52**<br>**[0.43-0.63]**<br>**(2.0×10<sup>-11</sup>)** | **1.98**<br>**[1.36-2.89]**<br>**(4.0×10<sup>-4</sup>)** |

Associations between incident mortality risk, Covid-19 clusters identified and use of Hydroxychloroquine (HCQ), in the incremental models tested and in a sensitivity analysis including all the drugs used for Covid-19 treatment. Hazard Ratios with 95% confidence intervals (HR [CI]) and relevant p-values (in brackets) are reported. Significant HRs ( $p < 0.05$ ) are highlighted in bold.

### Associations with a combined Covid-19 outcome

When we modelled the risk of bad clinical outcomes of the disease – i.e. severe Covid-19 manifestations, access to intensive care unit or death – as a function of clusters and HCQ use, we observed results in line with survival analyses, with increased risk for cluster 2 and decreased risk for HCQ users, both in the additive and in the interactive model (Table 3). While the cluster*HCQ interaction showed only a trend of significance (p = 0.08), HCQ still presented a significant protective association in the low risk cluster (OR [CI] = 0.67 [0.56-0.79], p = 4.0×10^−6^) and a substantially null association in the high risk cluster (OR [CI] = 0.98 [0.66-1.46], p = 0.92).

**Table 3.** Results of logistic regressions modelling Covid-19 composite bad outcome risk.

| Model | N | Cluster 2 vs 1 | HCQ | Cluster*HCQ |
| --- | --- | --- | --- | --- |
| Bad outcome □ Cluster | 4,373 | **2.53**<br>[2.09-3.08]<br>($< 10^{-15}$) | - | - |
| Bad outcome □ Cluster + HCQ | 4,265 | **2.53**<br>[2.08-3.09]<br>($< 10^{-15}$) | **0.71**<br>[0.61-0.83]<br>($2.1 \times 10^{-5}$) | - |
| Bad outcome □ Cluster + HCQ +<br>Cluster*HCQ | 4,265 | **1.94**<br>[1.35-2.79]<br>($3.8 \times 10^{-4}$) | **0.67**<br>[0.56-0.79]<br>($4.0 \times 10^{-6}$) | 1.46<br>[0.95-2.26]<br>0.08 |
| Bad outcome □ Cluster + HCQ +<br>Cluster*HCQ + Other Drugs | 3,786 | **1.80**<br>[1.21-2.68]<br>($3.8 \times 10^{-3}$) | **0.55**<br>[0.45-0.66]<br>($9.0 \times 10^{-10}$) | 1.47<br>[0.92-2.36]<br>(0.10) |

The composite bad outcome was defined as one of the following: death, access to intensive care unit or severe Covid-19 manifestation (either severe pneumonia or ARDS). Associations were tested in three incremental models and in a sensitivity analysis including all the drugs used for Covid-19 treatment, as for Cox PH regressions. Odds Ratios with 95% confidence intervals (OR [CI]) and relevant p-values (in brackets) are reported. Significant ORs ( $p < 0.05$ ) are highlighted in bold.

## Discussion

In the present work, we report a hierarchical clustering analysis applied to patients hospitalized for SARS-CoV-2 infection, which revealed the existence of two separate clusters of Covid-19 patients, based on their clinical and socio-demographic characteristics: one of younger patients with less comorbidities, lower circulating inflammation and better renal function (“low risk” cluster), and one of older and more comorbid patients, more prevalently men and smokers (“high risk” cluster). The former cluster showed a higher prevalence of severe manifestations of Covid-19, ranging from severe pneumonia to ARDS. Moreover, survival analyses showed an almost four-fold increase of incident in-hospital mortality for the high risk compared to the low risk cluster. Although a previous study attempted to identify subtypes of Covid-19 patients, associating them with disease severity [33], this represents the first attempt to use clustering in disentangling the effect of HCQ on different types of patients, by testing associations with incident in-hospital mortality risk. Specifically, we tested and observed both additive and interactive associations of HCQ and Covid-19 subtypes. Indeed, the high risk cluster was consistently associated with increased mortality across all models, while treatment with HCQ was generally associated with a halving of death risk, in line with previous evidence from both observational [6–13] and intervention studies [19]. While we already reported evidence suggesting a protective influence of HCQ against mortality in a largely overlapping sample [13], here we have further deepened this relationship by testing and reporting a significant association between cluster-by-HCQ interaction and mortality, which was driven by a differential association within the two clusters. Indeed, the low risk cluster showed a significant “protective” influence of HCQ on in-hospital deaths, while the high risk cluster showed a concordant but non-significant association. This represents an element of novelty of the present study, since in our previous work we observed a “protective” association between HCQ and mortality within the totality of patients (about 75% of the current sample size), and when stratifying by age, sex and other characteristics [13], but not within different subtypes of patients combining all these characteristics together in a non-linear setting, as can be built through unsupervised ML algorithms. Moreover, here our evidence is supported also by concordant associations with a composite and possibly more robust outcome of the disease, based on the occurrence of death, access to ICU and severity of manifestations.

Recently, an approach based on the definition of subtypes of Covid-19 patients has been already proven to be successful in identifying which patients benefit most from HCQ treatment, in a multi-center trial involving six US hospitals and 290 patients hospitalized for Covid-19, the IDENTIFY study [19]. HCQ treatment was associated with higher survival in the treated harm, and especially within those patients which were predicted to benefit most based on a supervised ML algorithm applied to their characteristics, which included blood pressure, heart rate, temperature, respiratory rate, oxygen saturation, white blood cell and platelet count, lactate, blood urea nitrogen, creatinine, and bilirubin levels [19]. Interestingly, lactate and creatinine levels where the most important features in this algorithm [19], the latter representing an index of renal function, which was also a characteristic feature of the low risk cluster in the present study, where HCQ was more effective. Moreover, patients eligible for HCQ treatment as derived by the algorithm of (Burdick et al., 2020) showed to be younger and less comorbid than the whole population studied, in line with the evidence reported in the present work, suggesting that HCQ treatment may be more effective for younger patients with better general health conditions.

### Strengths and Limitations

Although to our knowledge this study represents the largest and broadest cluster analysis on Covid-19 patients and a novel approach in analyzing the influence of HCQ treatment on Covid-19 mortality and outcomes, it also presents few limitations.

First, the observational retrospective design does not allow us to completely control for confounders and randomization of treatments across individuals. The former aspect is quite unlikely since a potential residual confounder should be strongly associated with in-hospital mortality to take away observed associations in the interactive model, as suggested by the computed E-values [31,32]. As for drug therapy, we cannot rule out that assignment to specific treatments was driven also by clinical conditions of the participants, as usually found in common clinical practice. For the same reason, the protective association observed for HCQ may be hypothesized to be driven by other co-administered medications. However, here HCQ and patients’ cluster showed significant independent associations, which remained substantially stable across models and survived correction for other drugs in use for Covid-19 treatment. Last, our evidence is in contrast with RCTs published so far [14–18], which are commonly conceived as the gold standard for establishing drug efficacy and safety. While we generally agree with this view, we would like to underline that these studies did not randomize patients to treatment arms based on combinations of their features, but rather based on single characteristics such as age and sex. This may be the reason for this discrepancy, along with the hypothesis that the high dosage of HCQ administered in RCTs may be harmful for patients, compared to lower dosages reported in observational studies supporting HCQ efficacy [5]. Of interest, a recent critical review underlined the aspect of suboptimal randomization methods of RCTs, which often do not take into account the whole patient profile and disease severity and may lead to misleading conclusions [34].

### Conclusions

Overall, the evidence supported here and elsewhere [19] suggests that HCQ treatment may be more effective in specific subtypes of Covid-19 patients and indicates machine learning as a useful approach to identify the most “promising” patients in terms of success rate of this treatment. Ideally, a trial administering low dosages of HCQ (≤400 mg/day) and randomizing subjects based on their Covid-19 subtype profile rather than on single characteristics may be warranted to clarify the effects of HCQ on mortality risk in SARS-CoV-2 infection, especially within those patients with a “low risk” profile.

This may help solving current controversies on the use of HCQ as a medication for Covid-19 and maximize the efficacy of treatment strategies for this yet largely unknown disease, especially in low-income and developing countries with poorer national health systems.

## Data Availability

Raw data analyzed in the present work may be made available under approval by each local center involved in the study, in a way which does not affect patients' privacy.

## URLs

R: https://www.r-project.org/

Keras package: https://cran.r-project.org/web/packages/keras/index.html

VIM package: https://cran.r-project.org/web/packages/VIM/index.html

Cluster package: https://cran.rproject.org/web/packages/cluster/index.html

Factoextra package: https://cran.rproject.org/web/packages/factoextra/index.html

E-value calculator: https://www.evalue-calculator.com/

## Author contributions

ADiC, RDC and LI conceived the CORIST study. AG, ADiC and LI conceptualized the present work. AG performed statistical analyses and wrote the first draft of the manuscript, under the supervision of ADiC and LI. All the co-authors contributed to collection, curation and elaboration of the analyzed data, and/or to a critical review and editing of the manuscript.

## Acknowledgements

We thank the participating clinical centers included in this cohort. This Article is dedicated to all the patients who suffered or died, often in solitude, due to COVID-19; their tragic fate gave us moral strength to start and carry out this project. The Authors are responsible for the views expressed in this Article, which do not necessarily represent the views, decisions, or policies of the Institutions they are affiliated with.

*The COVID-19 RISK and Treatments (CORIST) Collaboration*: Augusto Di Castelnuovo1^, Alessandro Gialluisi2^, Andrea Antinori3, Nausicaa Berselli4, Lorenzo Blandi5, Marialaura Bonaccio2, Raffaele Bruno6,7, Roberto Cauda8,9, Simona Costanzo2, Giovanni Guaraldi10, Lorenzo Menicanti11, Marco Mennuni12, Ilaria My13, Giustino Parruti14, Giuseppe Patti12, Stefano Perlini15,16, Francesca Santilli17, Carlo Signorelli18, Giulio Stefanini13, Alessandra Vergori19, Walter Ageno20, Antonella Agodi21, Piergiuseppe Agostoni22,23, Luca Aiello24, Samir Al Moghazi25, Rosa Arboretti26, Filippo Aucella27, Greta Barbieri28, Martina Barchitta29, Paolo Bonfanti30,31, Francesco Cacciatore32, Lucia Caiano20, Francesco Cannata13, Laura Carrozzi33, Antonio Cascio34, Giacomo Castiglione35, Arturo Cicullo8, Antonella Cingolani8,9, Francesco Cipollone17, Claudia Colomba34, Crizia Colombo12, Annalisa Crisetti27, Francesca Crosta14, Gian Battista Danzi36, Damiano D’Ardes17, Katleen de Gaetano Donati 8,9, Francesco Di Gennaro37, Giuseppe Di Tano36, Gianpiero D’Offizi38, Francesco Maria Fusco39, Carlo Gaudiosi40, Ivan Gentile41, Francesco Gianfagna1,20, Gabriele Giuliano8, Emauele Graziani42, Gabriella Guarnieri43, Valerio Langella44, Giovanni Larizza45, Armando Leone46, Gloria Maccagni36, Federica Magni20, Stefano Maitan24, Sandro Mancarella47, Rosa Manuele48, Massimo Mapelli22,23, Riccardo Maragna22,23, Rossella Marcucci49, Giulio Maresca44, Silvia Marongiu50, Claudia Marotta37, Lorenzo Marra46, Franco Mastroianni45, Alessandro Mengozzi51, Marianna Meschiari10, Jovana Milic10, Filippo Minutolo52, Roberta Mussinelli16, Cristina Mussini10, Maria Musso53, Anna Odone5, Marco Olivieri54, Antonella Palimodde50, Emanuela Pasi42, Raffaele Pesavento55, Francesco Petri30, Carlo A Pivato13, Venerino Poletti56,57, Claudia Ravaglia56, Giulia Righetti45, Andrea Rognoni12, Marco Rossato55, Ilaria Rossi17, Marianna Rossi30, Anna Sabena15, Francesco Salinaro15, Vincenzo Sangiovanni39, Carlo Sanrocco14, Nicola Schiano Moriello41, Laura Scorzolini58, Raffaella Sgariglia47, Paola Giustina Simeone14, Michele Spinicci49, Enrica Tamburrini8, Carlo Torti59, Enrico Maria Trecarichi59, Roberto Vettor55, Andrea Vianello43, Marco Vinceti4,60, Agostino Virdis51, Raffaele De Caterina33, Licia Iacoviello2,20#

^Equal contribution; #Correspondence to: Licia Iacoviello, MD, PhD; Department of Epidemiology and Prevention, IRCCS Neuromed, Via dell’Elettronica, 86077 Pozzilli (IS), Italy; Phone: +39-3485108779. Affiliations are reported in the Supplementary File.

