## Supplementary File for "Disentangling the association of hydroxychloroquine treatment with mortality in Covid-19 hospitalized patients through Hierarchical Clustering"

**Supplementary Methods**

**Figure S1. Cross-patient dissimilarity matrix based on Gower distance.**

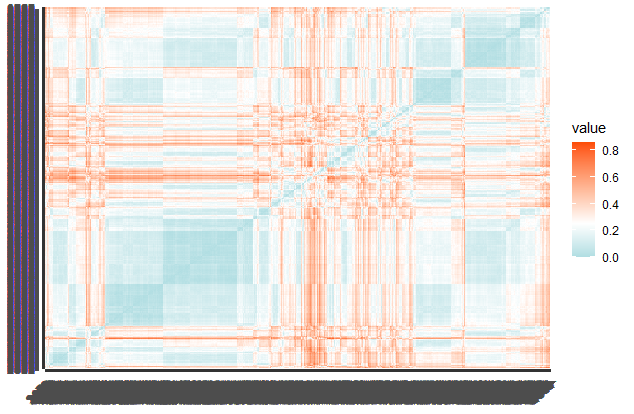

Each squared dot represents a pairwise comparison between patients.

**Figure S2.** Optimal number of clusters (k) to apply in the hierarchical clustering analysis.

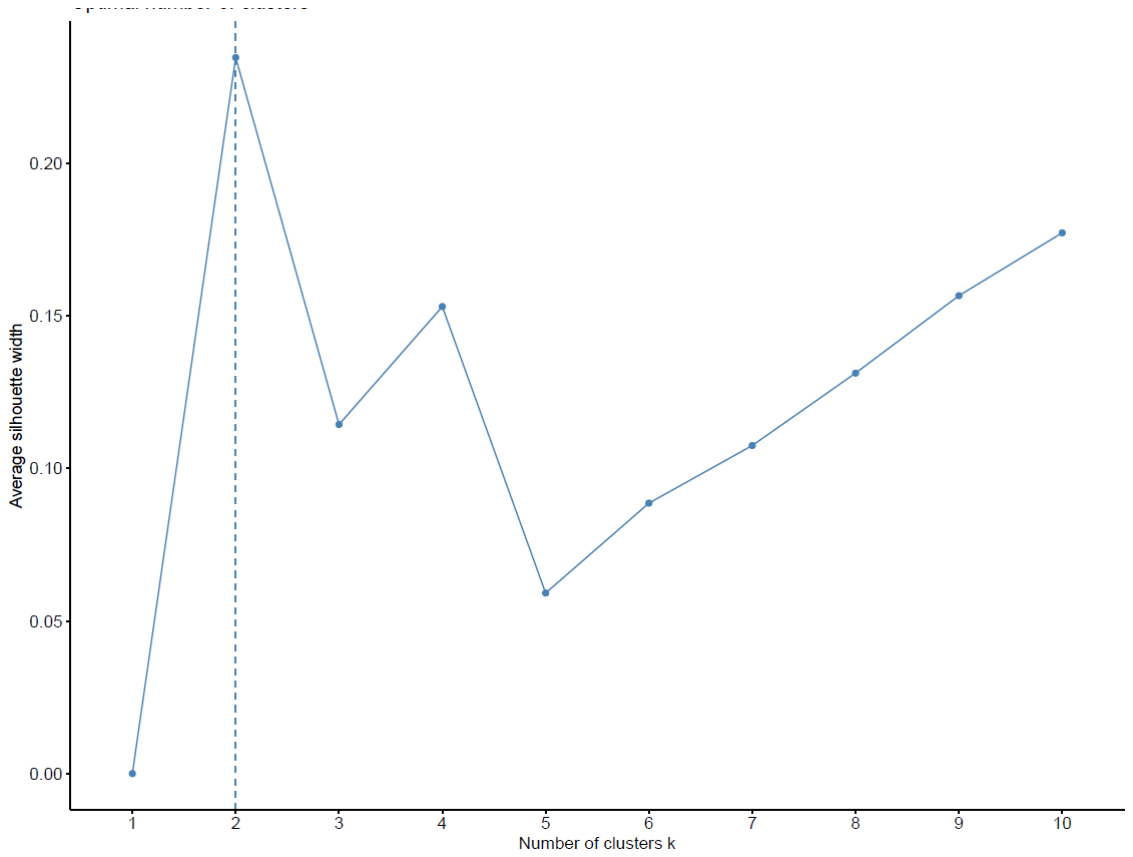

The optimal number of clusters (k) was computed through the Silhouette method. The high value of the “Average silhouette width” (> 0.2) indicates that each element is well matched to its cluster and poorly matched to neighboring clusters. The highest peak in the plot is taken as the optimal number of clusters (k=2, in this case).

**Figure S3.** Check for basic assumptions of Cox PH models.

**a)**

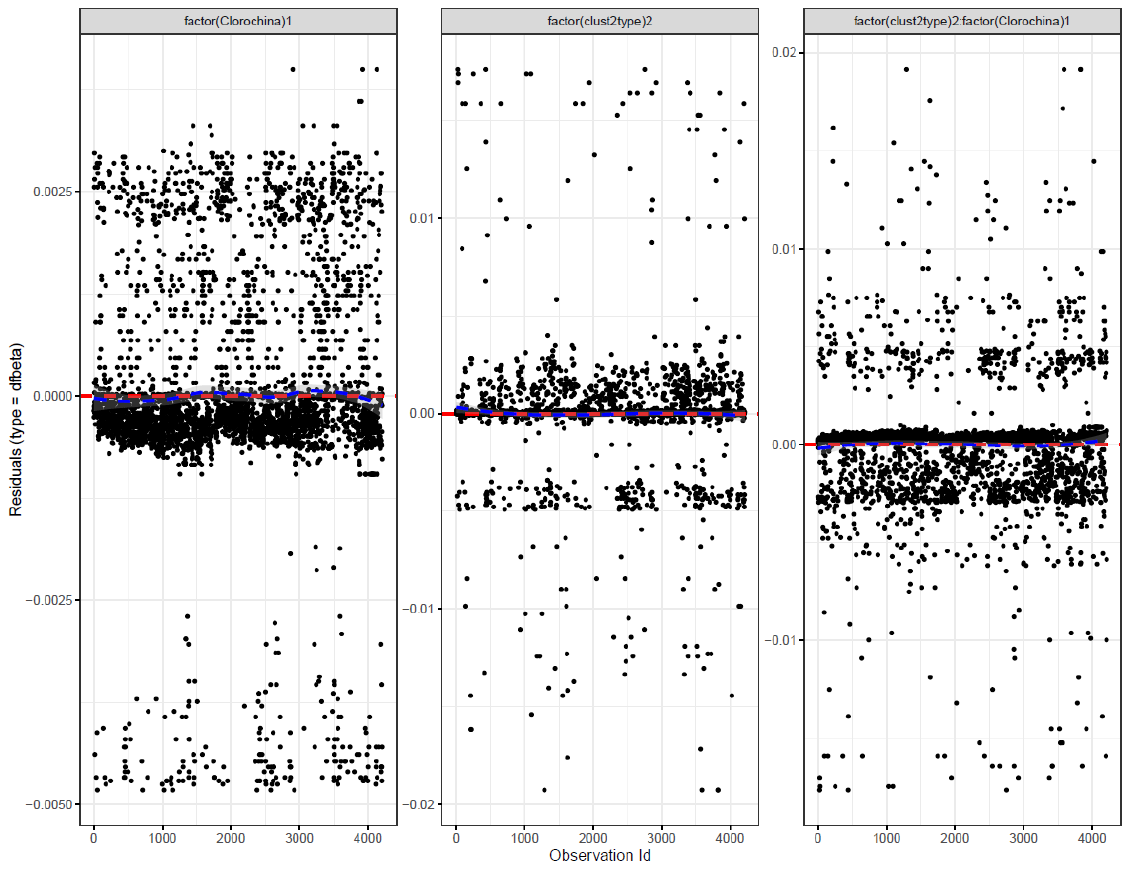

**b)**

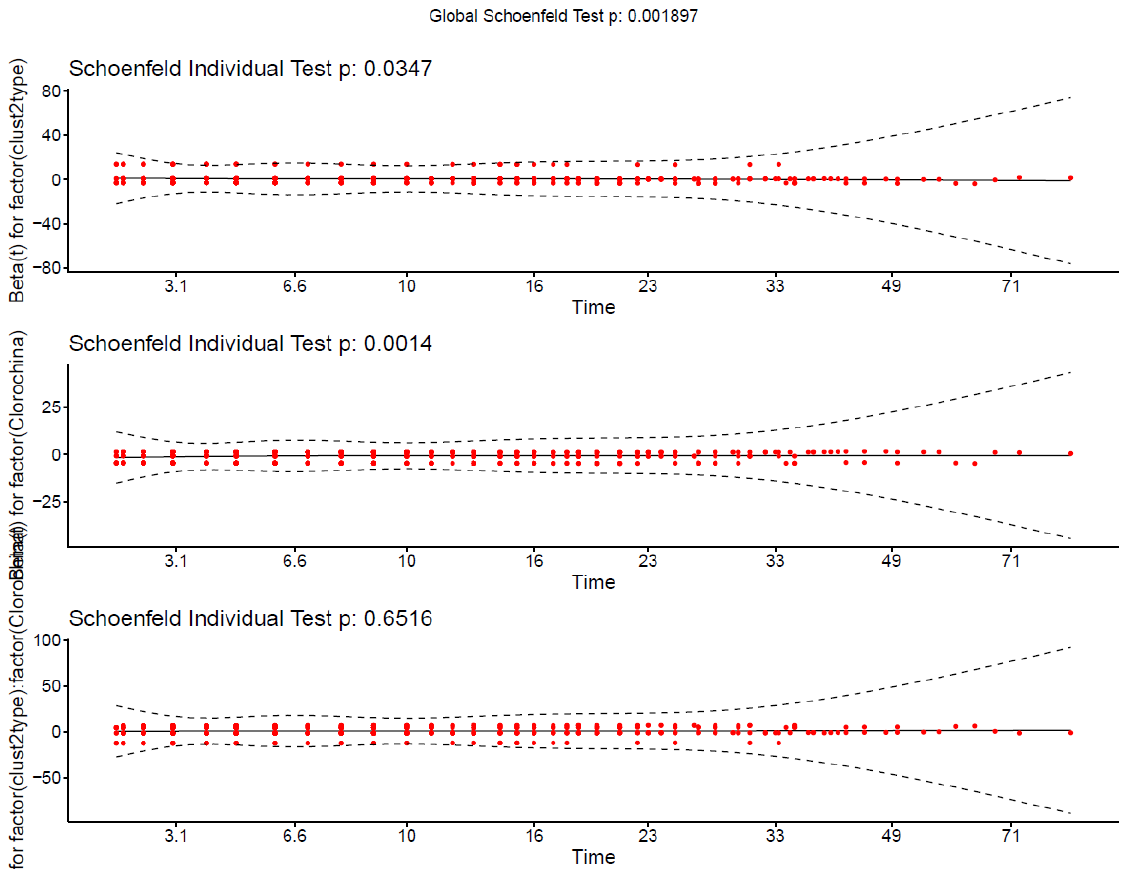

Test for **a)** lack of influential observations (using dfbeta residuals) and **b)** proportionality of hazards assumptions (Schoenfeld residuals) are reported. No dfbeta residuals >2/sqrt(N) were observed, suggesting the absence of outlier observations. Although no anomaly in the trends of Schoenfeld residuals vs follow-up time was observed, we detected statistical evidence for violations of the PH assumption, which prompted us to perform analyses also including an interaction term with time (see below).

**Supplementary Results**

**Table S1.** Contingency table of Covid-19 cluster by disease severity.

| Covid-19 disease  severity vs clusters | | Patients’ cluster | |
| --- | --- | --- | --- |
|  |  | Cluster 1 – low risk | Cluster 2 – high risk |
| Disease severity | Asymptomatic/mild | 524 (13.5%) | 39 (8.2%) |
|  | Non-severe pneumonia | 1577 (40.6%) | 124 (26.0%) |
|  | Severe pneumonia | 996 (25.7%) | 200 (41.9%) |
|  | ARDS | 784 (20.2%) | 114 (23.9%) |
| Total N (with available classification) | | 3,881 | 477 |

Only participants with disease severity classification available were included in the table.

Abbreviations: ARDS = Acute respiratory distress syndrome.

**Table S2.** Contingency tables of Covid-19 clusters by drugs used for treatment.

| **Drug** | **Use** | **Cluster 1**  **low risk** | **Cluster 2**  **high risk** | **Fisher Exact Test (p)** |
| --- | --- | --- | --- | --- |
| **Hydroxychloroquine** | **No** | **861 (22.6%)** | **139 (30.1%)** | **4.6×10^-4^** |
|  | **Yes** | **2,948 (77.4%)** | **322 (69.9%)** |  |
| **Anti-hypertensive drugs** | **No** | **2,464 (63.9%)** | **136 (28.7%)** | **<10^-15^** |
|  | **Yes** | **1,391 (36.1%)** | **337 (71.3%)** |  |
| Anti-interleukin-6 antibody | No | 3,218 (85.0%) | 398 (84.9%) | 0.95 |
|  | Yes | 568 (15.0%) | 71 (15.1%) |  |
| Remdesivir | No | 3,731 (97.4%) | 449 (96.5%) | 0.29 |
|  | Yes | 99 (2.6%) | 16 (3.5%) |  |
| **Lopinavir/Darunavir** | **No** | **2,015 (53.6%)** | **271 (58.8%)** | **0.04** |
|  | **Yes** | **1,746 (46.4%)** | **190 (41.2%)** |  |
| **Corticosteroids** | **No** | **2,286 (65.6%)** | **250 (56.3%)** | **1.5×10^-4^** |
|  | **Yes** | **1,199 (34.4%)** | **194 (43.7%)** |  |

Absolute count of drug users and relative frequency within each cluster are reported. Significant comparisons (p < 0.05) are highlighted in bold. Only participants with medication information available were included in each table.

**Table S3.** Results of Cox PH regressions modelling incident mortality risk, including interaction terms with time.

| **Model** | **Cluster**  **2 vs 1** | **HCQ**  **Yes vs no** | **Cluster*HCQ** |
| --- | --- | --- | --- |
| Death ⁓ Cluster | **3.71**  **[3.15-4.38]** | **-** | **-** |
| Death ⁓ Cluster + HCQ | **3.93**  **[3.31-4.66]** | **0.69**  **[0.59-0.81]** | **-** |
| Death ⁓ Cluster + HCQ + Cluster*HCQ | **2.77**  **[2.33-3.28]** | **0.59**  **[0.50-0.69]** | **1.81**  **[1.49-2.20]** |
| Death ⁓ HCQ (within “low risk” cluster) | **-** | **0.61**  **[0.51-0.72]** | **-** |
| Death ⁓ HCQ (within “high risk” cluster) | **-** | 1.06  [0.77-1.48] | **-** |

**Associations between incident mortality risk, Covid-19 clusters identified and use of Hydroxychloroquine (HCQ), in the three incremental models tested in the total sample, as well as within each cluster. Hazard Ratios** with 95% confidence intervals (HR [CI]) are reported. **Significant HRs (p < 0.05) are highlighted in bold.**

**The COVID-19 RISK and Treatments (CORIST) Collaboration:**

**Authors list**

Augusto Di Castelnuovo1^, Alessandro Gialluisi2^, Andrea Antinori3, Nausicaa Berselli4, Lorenzo Blandi5, Marialaura Bonaccio2, Raffaele Bruno6,7, Roberto Cauda8,9, Simona Costanzo2, Giovanni Guaraldi10, Lorenzo Menicanti11, Marco Mennuni12, Ilaria My13, Giustino Parruti14, Giuseppe Patti12, Stefano Perlini15,16, Francesca Santilli17, Carlo Signorelli18, Giulio Stefanini13, Alessandra Vergori19, Walter Ageno20, Antonella Agodi21, Piergiuseppe Agostoni22,23, Luca Aiello24, Samir Al Moghazi25, Rosa Arboretti26, Filippo Aucella27, Greta Barbieri28, Martina Barchitta29, Paolo Bonfanti30,31, Francesco Cacciatore32, Lucia Caiano20, Francesco Cannata13, Laura Carrozzi33, Antonio Cascio34, Giacomo Castiglione35, Arturo Cicullo8, Antonella Cingolani8,9, Francesco Cipollone17, Claudia Colomba34, Crizia Colombo12, Annalisa Crisetti27, Francesca Crosta14, Gian Battista Danzi36, Damiano D'Ardes17, Katleen de Gaetano Donati	8,9, Francesco Di Gennaro37, Giuseppe Di Tano36, Gianpiero D'Offizi38, Francesco Maria Fusco39, Carlo Gaudiosi40, Ivan Gentile41, Francesco Gianfagna1,20, Gabriele Giuliano8, Emauele Graziani42, Gabriella Guarnieri43, Valerio Langella44, Giovanni Larizza45, Armando Leone46, Gloria Maccagni36, Federica Magni20, Stefano Maitan24, Sandro Mancarella47, Rosa Manuele48, Massimo Mapelli22,23, Riccardo Maragna22,23, Rossella Marcucci49, Giulio Maresca44, Silvia Marongiu50, Claudia Marotta37, Lorenzo Marra46, Franco Mastroianni45, Alessandro Mengozzi51, Marianna Meschiari10, Jovana Milic10, Filippo Minutolo52, Roberta Mussinelli16, Cristina Mussini10, Maria Musso53, Anna Odone5, Marco Olivieri54, Antonella Palimodde50, Emanuela Pasi42, Raffaele Pesavento55, Francesco Petri30, Carlo A Pivato13, Venerino Poletti56,57, Claudia Ravaglia56, Giulia Righetti45, Andrea Rognoni12, Marco Rossato55, Ilaria Rossi17, Marianna Rossi30, Anna Sabena15, Francesco Salinaro15, Vincenzo Sangiovanni39, Carlo Sanrocco14, Nicola Schiano Moriello41, Laura Scorzolini58, Raffaella Sgariglia47, Paola Giustina Simeone14, Michele Spinicci49, Enrica Tamburrini8, Carlo Torti59, Enrico Maria Trecarichi59, Roberto Vettor55, Andrea Vianello43, Marco Vinceti4,60, Agostino Virdis51, Raffaele De Caterina33, Licia Iacoviello2,20#

^Equal contribution

#Correspondence to: Licia Iacoviello, MD, PhD; Department of Epidemiology and Prevention, IRCCS Neuromed, Via dell'Elettronica, 86077 Pozzilli (IS), Italy; Phone: +39-3485108779;.

**Affiliations**

1	Mediterranea Cardiocentro, Napoli. Italy

2	Department of Epidemiology and Prevention, IRCCS Neuromed, Pozzilli (IS). Italy

3	UOC Immunodeficienze Virali, National Institute for Infectious Diseases “L. Spallanzani”, IRCCS, Rome Italy

4	Section of Public Health, Department of Biomedical, Metabolic and Neural Sciences, University of Modena and Reggio Emilia, Modena, Italy;

5	Università di Pavia, Pavia, Italy

6	Division of Infectious Diseases I, Fondazione IRCCS Policlinico San Matteo, Pavia. Italy

7	Department of Clinical, Surgical, Diagnostic, and Paediatric Sciences, University of Pavia, Pavia. Italy

8	Fondazione Policlinico Universitario A. Gemelli IRCCS, Roma. Italy

9	Università Cattolica del Sacro Cuore- Dipartimento di Sicurezza e Bioetica Sede di Roma, Italy

10	Infectious Disease Unit, Department of Surgical, Medical, Dental and Morphological Sciences, University of Modena and Reggio Emilia, Modena, Italy

11	IRCCS Policlinico San Donato, San Donato Milanese. Italy

12	University of Eastern Piedmont, Maggiore della Carità Hospital, Novara. Italy

13	Humanitas Clinical and Research Hospital IRCCS, Rozzano-Milano. Italy

14	Department of Infectious Disease, Azienda Sanitaria Locale (AUSL) di Pescara, Pescara, Italy;

15	Emergency Department, IRCCS Policlinico San Matteo Foundation, Pavia. Italy

16	Department of Internal Medicine, University of Pavia, Pavia. Italy

17	Department of Medicine and Aging, Clinica Medica, "SS. Annunziata" Hospital and University of Chieti, Chieti. Italy

18	School of Medicine, Vita-Salute San Raffaele University, Milano. Italy

19	HIV/AIDS Department, National Institute for Infectious Diseases "Lazzaro Spallanzani"-IRCCS, Roma, Italy

20	Department of Medicine and Surgery, University of Insubria, Varese. Italy

21	Department of Medical and Surgical Sciences and Advanced Technologies "G.F. Ingrassia", University of Catania; AOU Policlinico "G.Rodolico - San Marco", Catania. Italy

22	Centro Cardiologico Monzino IRCCS, Milano. Italy

23	Department of Clinical Sciences and Community Health, Cardiovascular Section, University of Milano

24	UOC. Anestesia e Rianimazione. Dipartimento di Chirurgia Generale Ospedale Morgagni-Pierantoni Forlì, Italy

25	UOC Infezioni Sistemiche dell'Immunodepresso, National Institute for Infectious Diseases L. Spallanzani, IRCCS, Rome Italy

26	Department of Civil Environmental and Architectural Engineering. University of Padova. Padova. Italy

27	Fondazione I.R.C.C.S “Casa Sollievo della Sofferenza”, San Giovanni Rotondo, Foggia. Italy

28	Department of Surgical Medical and Molecular Medicine and Critical Care. Azienda Ospedaliera Universitaria Pisana, and University of Pisa, Pisa. Italy

29	Department of Medical and Surgical Sciences and Advanced Technologies "G.F. Ingrassia", University of Catania, Catania, Italy

30	UOC Malattie Infettive, Ospedale San Gerardo, ASST Monza, Monza. Italy

31	School of Medicine and Surgery, University of Milano-Bicocca, Milano. Italy

32	Department of Translational Medical Sciences. University of Naples, Federico II,Naples, Italy

33	Cardiovascular and Thoracic Department, Azienda Ospedaliero-Universitaria Pisana, and University of Pisa, Pisa, Italy

34	Infectious and Tropical Diseases Unit- Department of Health Promotion, Mother and Child Care, Internal Medicine and Medical Specialties (PROMISE) - University of Palermo, Palermo. Italy

35	Servizio di Anestesia e Rianimazione II UO Rianimazione Ospedale San Marco, AOU Policlinico "G. Rodolico - San Marco", Catania, Italy

36	Department of Cardiology, Ospedale di Cremona, Cremona, Ital

37	Medical Direction, IRCCS Neuromed, Pozzilli (IS). Italy

38	UOC Malattie Infettive-Epatologia. National Institute for Infectious Diseases L. Spallanzani. IRCCS. Roma. Italy

39	UOC Infezioni Sistemiche e dell’Immunodepresso, Azienda Ospedaliera dei Colli, Ospedale Cotugno, Napoli. Italy;

40	ASL Napoli3 Sud COVID HOSPITAL Boscotrecase (NA), Italy

41	Department of Clinical Medicine and Surgery. University of Naples "Federico II". Napoli. Italy

42	Medicina Interna. Ospedale di Ravenna. AUSL della Romagna, Ravenna. Italy

43	Respiratory Pathophysiology Division, Department of Cardiologic, Thoracic and Vascular Sciences, University of Padova, Padova, Italy

44	UOC Medicina COVID- PO S. Maria di Loreto Nuovo, ASL Na 1 Centro, Napoli, Italy

45	COVID-19 Unit. EE Ospedale Regionale F. Miulli, Acquaviva delle Fonti (BA). Italy

46	UOC di Pneumologia, P.O. San Giuseppe Moscati, Taranto. Italy

47	ASST Milano Nord - Ospedale Edoardo Bassini; Cinisello Balsamo, Italy

48	U.O. C. Malattie Infettive e Tropicali, P.O. "San Marco", AOU Policlinico "G. Rodolico - San Marco", Catania, Italy

49	Department of Experimental and Clinical Medicine. University of Florence and Azienda Ospedaliero-Universitaria Careggi. Firenze. Italy

50	P.O. Santissima Trinità di Cagliari. Cagliari. Italy

51	Department of Clinical and Experimental Medicine. Azienda Ospedaliera Universitaria Pisana. University of Pisa, Pisa. Italy

52	Dipartimento di Farmacia. Università di Pisa. Pisa. Italy.

53	UOC Malattie Infettive-Apparato Respiratorio, National Institute for Infectious Diseases “L. Spallanzani”, IRCCS, Rome Italy

54	Computer Service, University of Molise, Campobasso, Italy.

55	Clinica Medica 3. Department of Medicine - DIMED. University hospital of Padova. Padova. Italy

56	UOC Pneumologia. Dipartimento di Malattie Apparato Respiratorio e Torace. Ospedale Morgagni-Pierantoni Forlì, Forlì. Italy

57	Department of Respiratory Diseases & Allergy Aarhus University Hospital. Aarhus. Denmark

58	UOC Malattie Infettive ad Alta Intensità di Cura, National Institute for Infectious Diseases “L. Spallanzani”, IRCCS, Rome Italy

59	Infectious and Tropical Diseases Unit. Deparment of Medical and Surgical Sciences “Magna Graecia” University,Catanzaro. Italy

60	Department of Epidemiology, Boston University School of Public Health, Boston. USA
